# Creating an Early Diagnostic Method for Glaucoma Using Convolutional Neural Networks

**DOI:** 10.1101/2024.03.14.24304273

**Authors:** Areej A. Alqarni, Sanad H. Al Harbi, Irshad A. Subhan

## Abstract

According to the World Health Organization, glaucoma is a leading cause of blindness, accounting for over 12% of global blindness as it affects one in every 100 people. In fact, 79.6 million people worldwide live with blindness caused by glaucoma. This is because the current method for diagnosing glaucoma is by examining retinal fundus images. However, it is considerably difficult to distinguish the lesions’ features solely through manual observations by ophthalmologists, especially in the early phases. This study introduces a novel glaucoma detection method using attention-enhanced convolutional neural networks, achieving 98.9% accuracy and a swift 30-second detection time, vastly surpassing traditional diagnostic methods. The attention mechanism is utilized to learn pixel-wise features for accurate prediction. Several attention strategies have been developed to guide the networks in learning the important features and factors that affect localization accuracy. The algorithms were trained for glaucoma detection using Python 2.7, TensorFlow, Py Torch, and Keras Machine Learning-Based Applications. The methods were evaluated on Drishti-GS and RIM-ONE datasets with 361 training and 225 test sets, consisting of 344 healthy and 242 glaucomatous images. The proposed algorithms can achieve impressive results that show an increase in overall diagnostic efficiency, as the algorithm displays a 30-second detection time with 98.9% accuracy compared to the 72.3% accuracy of traditional testing methods. Finally, this algorithm has been implemented as a webpage, allowing patients to test for glaucoma. This webpage offers various services such as: connecting the patient to the nearest care setup; offering scientific articles regarding glaucoma; and a video game that supports eye-treatment yogic exercises to strengthen vision and focus. This early diagnostic method has the near future potential to decrease the percentage of irreversible vision loss due to glaucoma by 42.79% (the percentage was calculated using the mean absolute error function), which could prevent glaucoma from remaining the leading cause of blindness worldwide. Our glaucoma diagnostic webpage can be found at: [Glaucoma Detector](http://glaucomadiagnosis.com/) (glaucomadiagnosis.com)

## 1. Introduction

Glaucoma is a progressive eye disease that is among the leading causes of irreversible blindness. According to the World Health Organization, it causes over 12% of global blindness [1]. In 2010, it was estimated that 79.6 million people were diagnosed with glaucoma, a figure expected to increase to 111 million by the year 2040. The disordered physiological processes associated with this disease are multifactorial. However, the causes of glaucoma are usually associated with the build-up or intraocular pressure in the eye that results from blockage of intraocular fluid drainage [2]. The increased intraocular pressure damages the optic nerve that carries visual information of photoreceptors from the eye to the brain. Glaucoma is commonly referred to as the “hushed burglar of vision” since the early-stage symptoms are not explicitly defined and are difficult to quantify [3-8]. Glaucoma generally shows no signs until it has progressed to an advanced stage, at which point the damage becomes irrevocable. It has been reported that the damage to optic nerve fibers becomes noticeable and a reduction in the visual field is detected when about 40% of axons are already lost. (Figure 1). Thus, it is necessary to detect glaucoma at an early stage due to the fact that there are no perceptible indications in its preliminary stages. The condition is regarded as severe since the damage it causes is irredeemable, leading to perpetual vision loss if not promptly treated. While there is no prophylactic treatment for glaucoma, it is possible to avoid blindness by detecting and curing the condition at an initial stage [9-15]. Glaucoma is normally diagnosed by measures such as: obtaining a medical history of the patient; measuring intraocular pressure; performing a visual field loss test; and conducting a manual assessment of the optic disc using ophthalmoscopy to examine the shape and color of the optic nerve. The optic disc is the cross-sectional view of the optic nerve connecting to the retina of each eye. In retinal fundus images, it looks like a bright round spot. However, in cases of glaucoma, the intraocular pressure damages the nerve fibers constituting the optic nerve [15]. The boundary of the disc also dilates and the color changes from healthy pink to pale. (Figure 2).

**Figure 1.**
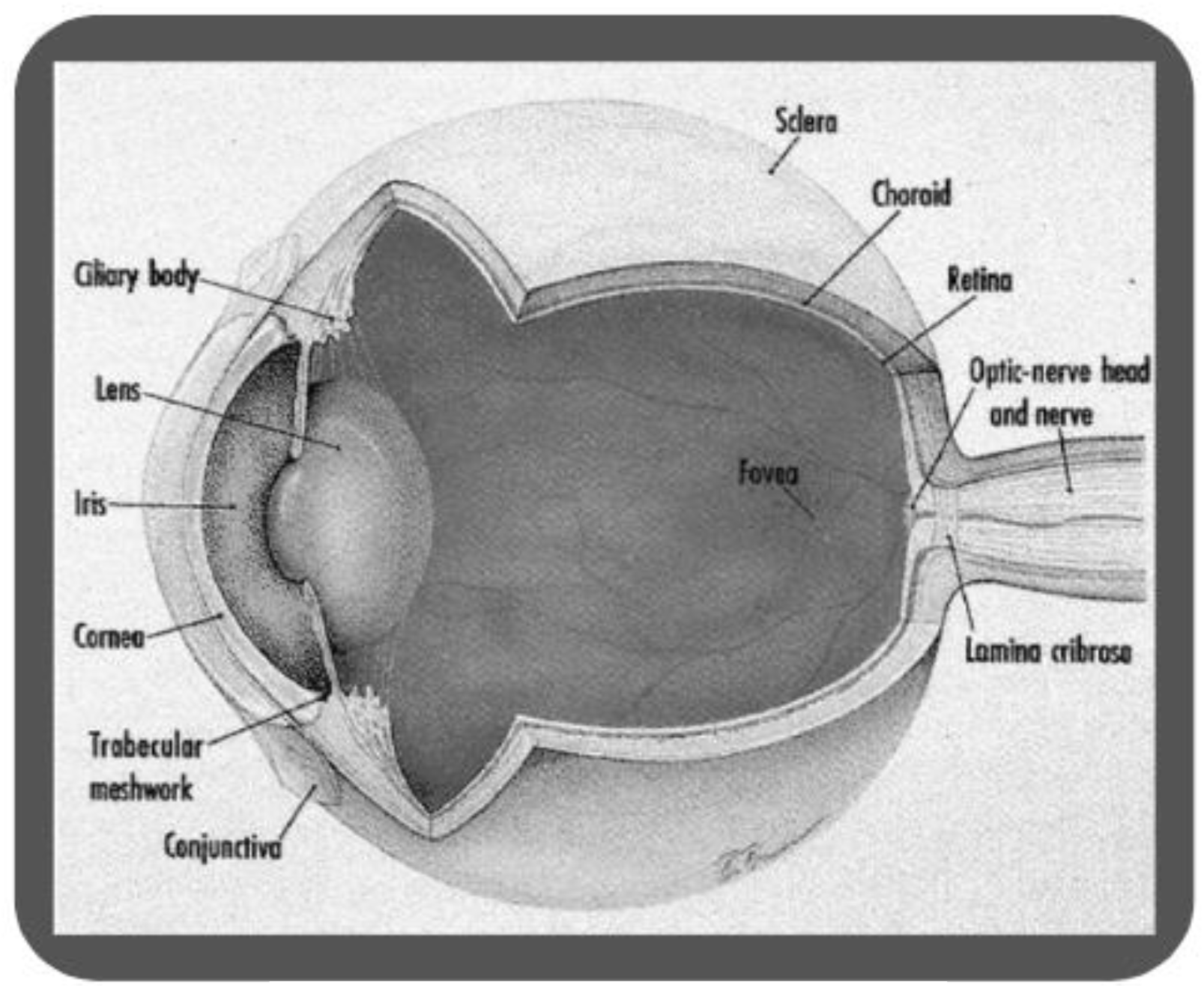
The anatomy of the human eye

**Figure 2.**
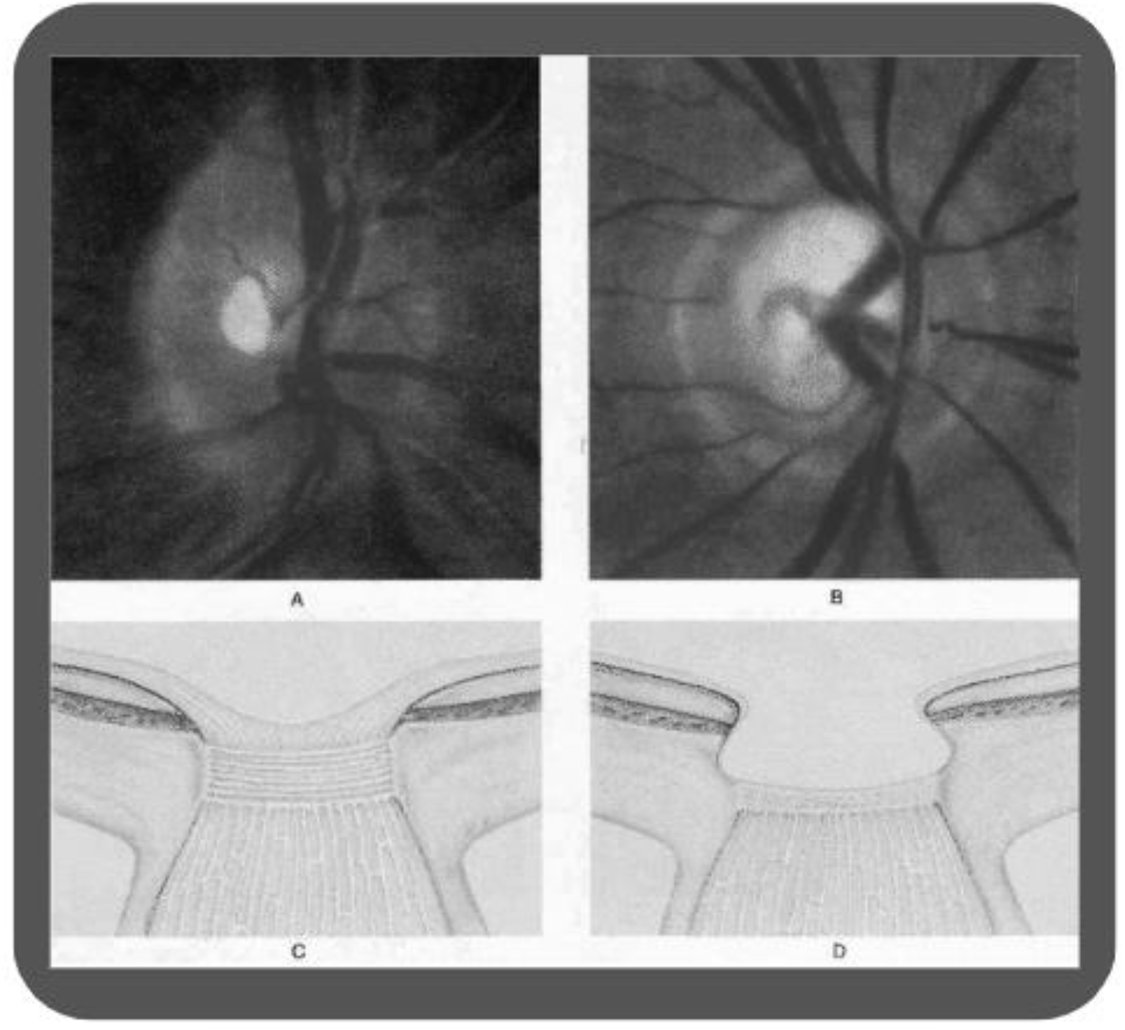
Glaucomatous and normal optic discs

**Table 1.**
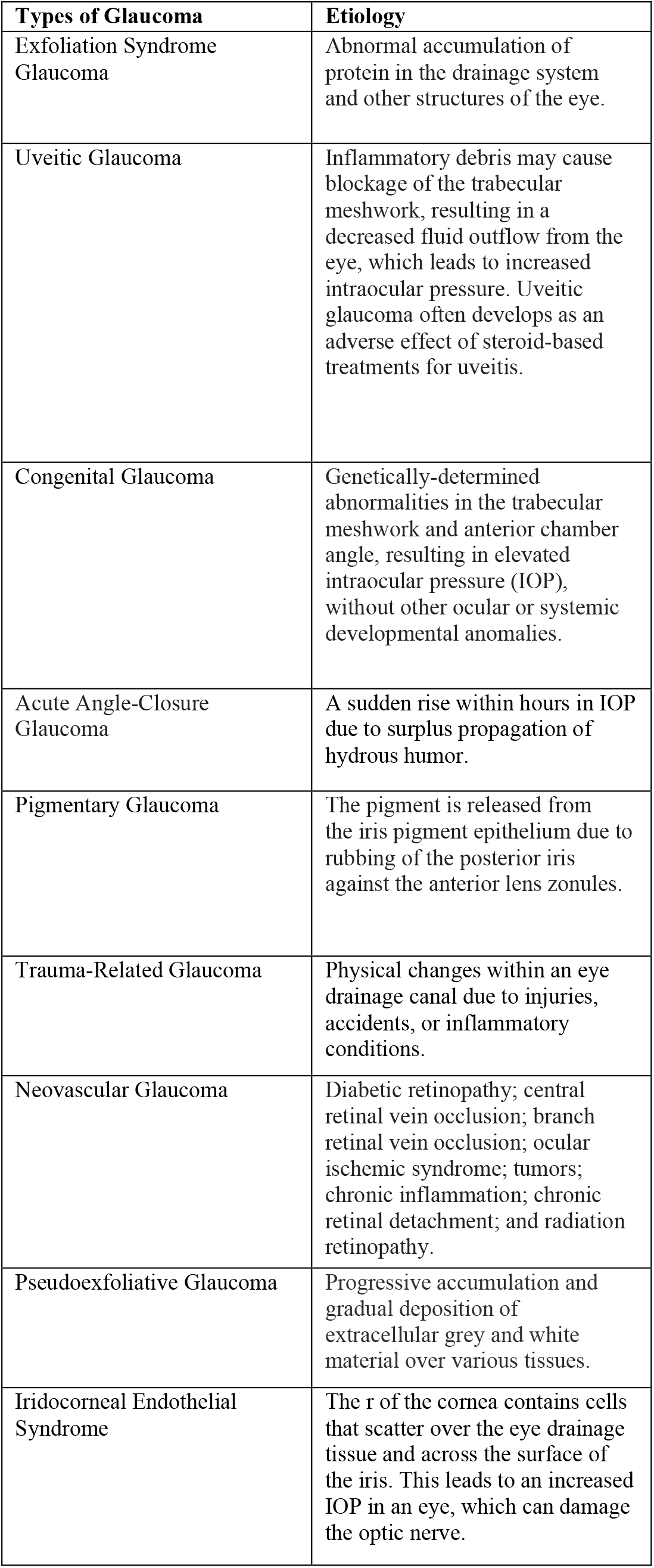
Types of Glaucoma.

All these are computed using the vertical disc and cup heights. Automatic methods thus generally rely on analysis of the optic disc region for glaucoma diagnosis [16-19]. Three types of glaucoma detection algorithms are commonly presented in the literature: structural, generic, and hybrid. The performance of structural glaucoma detection algorithms is thus highly dependent on the segmentation accuracy of the optic disc and cup from retinal images. Consequently, the accurate detection of the boundaries of both the disc and cup is essential for ensuring the efficiency of these methods [20]. Generic methods are based on the intuition that the damage to the optic nerve that is associated with glaucoma affects the overall characteristics of the optic disc region. Generic methods thus rely on a combination of statistical and textural features, computed from either the spatial or wavelet domains of the image. Hybrid approaches combine both structural measures, requiring optic disc and cup segmentation along with generic textural and statistical features for glaucoma detection [21-24]. Thereby, segmentation of this region of interest is not only useful for a more precise clinical assessment by ophthalmologists but also helpful in training a computer-based automated method for classification. Automated glaucoma detection methods are very sensitive; even the smallest error can affect the accuracy of the detection. Detecting subtle deforms in the images is considerably difficult even for experts in the field. However, deep learning algorithms are able to identify otherwise unnoticeable characteristics, if sufficiently trained [25-29]. Since computeraided diagnosis helps in the rehabilitation of eyes affected by the disease, it can be helpful in providing cost-effective screening on a large scale. These automated techniques can reduce human error, therefore providing a highly accurate, accessible, and overall efficient performance. In this work, an early detection method for glaucoma using deep learning technologies, specifically convolutional neural networks (CNNs), is proposed. According to current scientific findings, there is no fully automated deep learning-based method of disc localization that gives accurate and precise results independent of the datasets. Additionally, many existing automated methods show low-level accuracy, to the point of accepting inefficient results where the intersection over union between actual and predicted locations is only greater than 0. To place this in perspective, current detection methods display a 72.3% accuracy [30-39]. This value is relatively low compared to the diagnostic approaches discussed in this paper. This approach, which also offers a higher detection speed at a 30-second rate, introduces a new state of the art and sets the bar for correct localization at IOU > 70. The automated algorithms are implemented as a webpage in order to offer timely service in remote areas. Furthermore, this method is generally regarded as being more accessible than approaches only available in tertiary care setups. The novel approach to glaucoma detection utilizing a deep learning framework based on convolutional neural networks (CNNs) with an attention mechanism, is designed to accurately discern pixel-level features critical for early-stage glaucoma identification. The proposed algorithm, rigorously trained on extensive datasets, achieves a diagnostic accuracy of 98.9% and operates with a detection time of 30 seconds. By deploying this model on a web-based platform, the study aims to enhance the availability of early diagnostic tools and potentially contribute to the reduction of glaucoma-related vision impairment.

## 2. Methods

### 2.1 VARIABLES

- Independent Variables: Artificial intelligence-based code - Images (Raw Data)
- Dependent Variables: Accuracy – Sensitivity – Specificity
- Control Variables: Programming Language Python – Keras Software – Py Torch – TensorFlow

### 2.2 DATASETS

#### RIM-ONE (Figure 4)

RIM-ONE is an open retinal fundus image dataset with accurate gold standards of the optic nerve head provided by different experts. It includes images from healthy eyes as well as from eyes with glaucoma at different stages. A variety of measurements by zones of the optic disc is also proposed for the purpose of validation. Three Spanish hospitals have contributed to the development of this dataset: Hospital Universitario de Canarias, Hospital Clínico San Carlos, and Hospital Universitario Miguel Servet. This dataset contains 485 stereo eye fundus images with a resolution of 2144 × 1424. Two sets of ground truths for the optic disc and optic cup are available. The first set is commonly used for training and testing. The second set, which acts as a “human” baseline, also uses DCSeg as a tool for optic disc and cup segmentation of stereo and monocular retinal fundus images.

#### Drishti-GS (Figure 4)

Drishti-GS is an online retinal fundus image dataset for glaucoma analysis and the study of optic nerve head segmentation. This dataset generates manual segmentation for optic disc and optic cup. It also provides CDR and labels for each image as glaucomatous or healthy. The dataset contains 101 eye fundus images with varying resolutions. It also uses different imaging modalities, such as optical coherence tomography, Heidelberg retina tomograph, and fundus imaging, to assess glaucoma. Aravind Hospital contributed to the development of this dataset.

**Table 2.** Overview of datasets used for evaluating methods.

| Dataset | Total Size | Healthy | Glaucomatous | Train | Validate | Test |
| --- | --- | --- | --- | --- | --- | --- |
| RIM-One | 485 | 313 | 172 | 296 | 21 | 168 |
| Drishti-GS | 101 | 31 | 70 | 48 | 04 | 49 |

### 2.3 PROCEDURES

Convolutional neural networks, a class of artificial neural networks, is applied to analyze visual imagery.

#### 2.3.1 Data Partitioning

The data partitioning stage, which is the first stage, was executed to divide the dataset into different training and test sets.

Specifically, 3/5 of the circumpapillary images constitute the training set while the test set is defined by 2/5 of the data.

#### 2.3.2 Segmentation

A Unet-like architecture was used to learn different pixel-level features. The U-net was modified to have multiple inputs so that the network can receive more original raw pixel information during training. In this way, the risk of overfitting was avoided, and the network’s learning capability was enhanced. Hence, with specific training, the technology was able to identify glaucoma-affected eyes (pixel-wise). (Figure 3).

**Figure 3.**
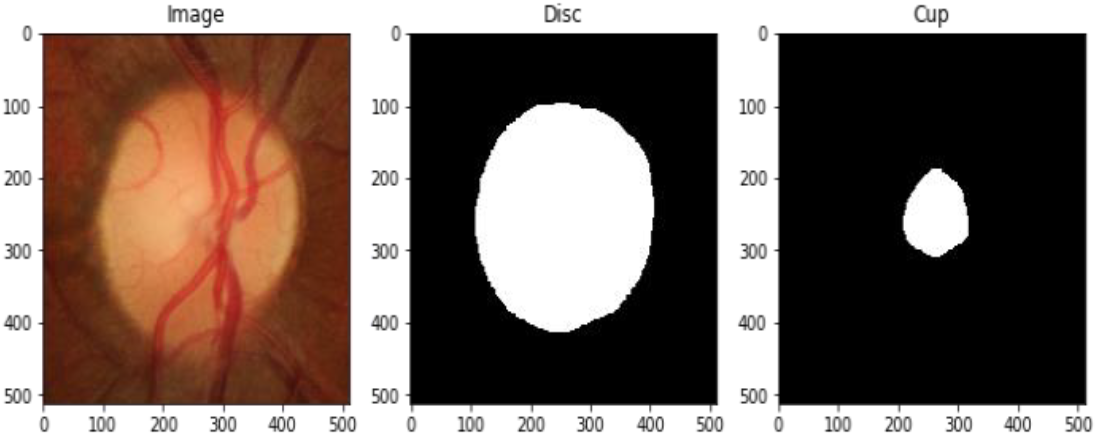
Optic cup and disc segmentation

**Figure 4.**
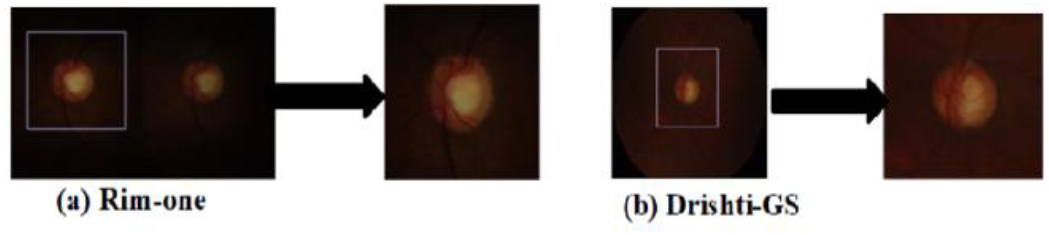
RIM-ONE and Drishti-GS datasets

#### 2.3.3 Regression

The segmentation task is more of an image regression instead of (a pixel classification problem), which deep learning (AI) usually needs in order to transform the low-level pixel information into high-level features. However, for the segmentation tasks, low-level pixel-wise features are more important. In contrast to learning to classify the pixels, directly mapping a retinal image to its corresponding label can keep more low-level pixel-wise features.

#### 2.3.4 Loss function

Due to the major pixel-wise similarities in training images, the mean absolute error (MAE) was adopted as the loss function in order to calculate the pixel-wise difference between the label and the prediction.

The MAE function is as follows:

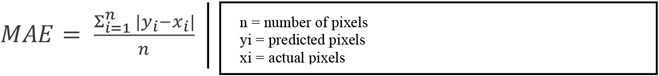

#### 2.3.5 Classification

After training the algorithm using segmentation (pixel by pixel), classification was used so that the technology could identify and distinguish glaucomatous eyes. In order to do so, the region and around-area of the optic cup/disc containing key pixel-wise features, such as the vertical disc diameter, the oval shape of the disc/cup, the ISNT rule and the yellow-orange rim, were used as they provide accurate results when distinguishing glaucoma.

##### Glaucoma Diagnosis Using CNNs versus a Traditional Method Comparative Study

Additionally, a comparative study was conducted to compare the CNN-based detection method with the traditional testing methods. used at King Abdullah Medical City in Saudi Arabia, and other tertiary care setups.

**Table 3.** CNN vs Traditional Methods Comparative Study.

| Approach | Time | Accuracy | Accessibility | MAE | Mean OC | Mean OD | Efficiency |
| --- | --- | --- | --- | --- | --- | --- | --- |
| CNN-based Method | 30-seconds to 1-minute | 98.9% | Accessible to patients worldwide (offering sufficient help to those in remote areas). | 0.0012. | 0.978. | 0.996. | Highly efficient, as it provides early, accessible, and accurate diagnosis. |
| Traditional Method | Varies for primary glaucoma. However, it takes approximately 20 to 45 minutes. | 86.2% | Most facilities are not available in primary or secondary hospitals: only in tertiary care setups. | 1.03. | 0.743. | 0.813. | Efficient. However, somewhat unreliable for early diagnosis as it is prone to be affected by clinician's bias or fatigue. It can also be inaccessible to certain demographics of people living in remote areas. |

#### 2.3.6 Data Prepossessing

First, the variance between training and validation images was reduced by cropping (600 × 600) the size of the ROI (region of interest) patches with the models that were already trained. This data processing is conducted to allow our model to focus on learning the necessary and important pixel-wise information needed for accurate prediction. (Figure 5).

**Figure 5.**
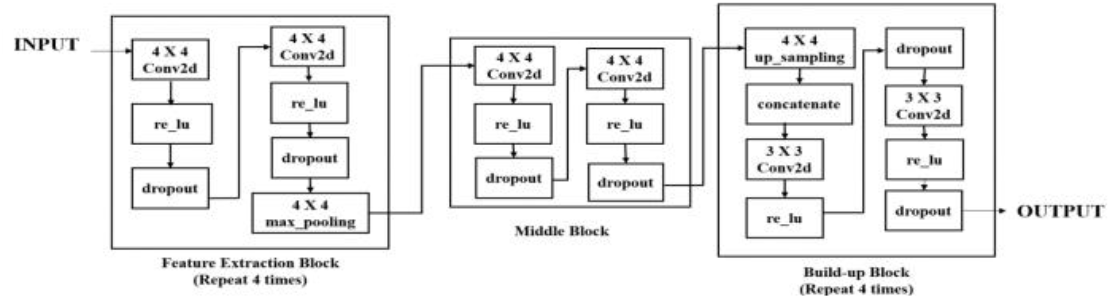
The flow of the proposed diagnostic method

#### 2.3.7 Data Augmentation Skills

such as image Rotation - 90/180/270 at various angles and image flipping, were also used in order to increase the number of training images. This increase is necessary as it is needed to ensure the network’s receptive field is sufficient and accurate. However, for the classification task, the ROIs were cropped for training and testing. The cropped regions were then resized to many different sizes for multiple deep learning training. Finally, the model was averaged as the final (most accurate) prediction. The training platforms used were: Python2.7, TensorFlow, Py Torch, and Keras.

## 3. Results

For segmentation, on the training set, the mean Optic Cup Dice is 0.900, the mean Optic Disc Dice is 0.937, and MAE CDR is 0.0012. On the validation set, mean Optic Cup Dice = 0.978, mean Optic Disc Dice = 0.996, and MAE CDR is 0.0043. On the testing set, mean Optic Cup Dice 0.989, mean Optic Disc Dice is 0.999, and MAE CDR is 0.0026 Best rank (results-online): 4th. For classification, on the training set AUC: is 1.0, and Sensitivity: is 1.0. On the validation set, AUC is 0.9803, and the Sensitivity: is 0.99. on the testing set, AUC: is 1.0, and Sensitivity: is 0.99 The latest (results-online) Rank: 1st. (Figure 6).

**Figure 6.**
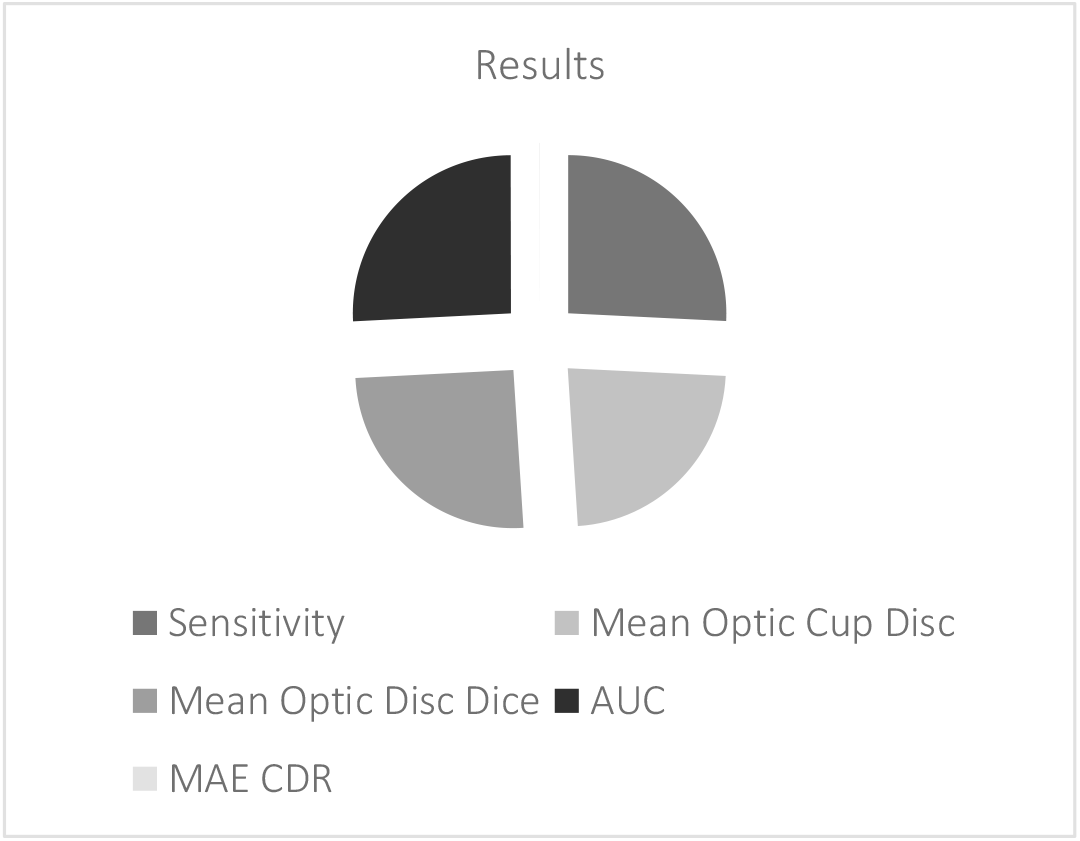
Results

### 2.3.8 Data Analysis

The data was assessed and validated for accuracy, sensitivity, and precision. The MAE function was used to assess potential errors. For automated localization of the optic disc, the model was trained for 100,000 iterations while the images were previously employed for training and validation. Once trained and evaluated on Drishti-GS and Rim-One datasets, the model was also tested on other publicly available databases and the results compared with some state-of-the-art methods developed specifically for those datasets. The results highlight the comparative performance of the fully automated method with state-of-the-art algorithms. The accuracy of the methods discussed in this paper are taken for 70% Intersection Over Union (IOU) The results reported by state-of-the-art algorithms are for IOU > 0. This diagnostic approach performed significantly better than existing methods, which means it was able to learn the discriminative representation of OD. It should be noted that most existing methods are normally designed with a particular dataset in focus whilst the algorithm that is proposed in this work was not designed to conform to any specific datasets. However, its performance is superior to those methods tailored specifically for those individual datasets. Thus, accuracy alone does not portray the true performance of the algorithm. Other performance metrics such as precision and sensitivity need to be assessed. Mathematical definitions of all these performances are given below. The results reveal a significant improvement in accuracy, sensitivity, and specificity levels of glaucoma diagnosis. In fact, traditional testing methods had a 72.3% accuracy, 73.1% sensitivity, and 82.1% specificity while this diagnostic approach showed a 98.9% accuracy, 98.8% sensitivity, and 99.1% specificity.

The mathematical definitions of accuracy, sensitivity, and specificity are as follows:

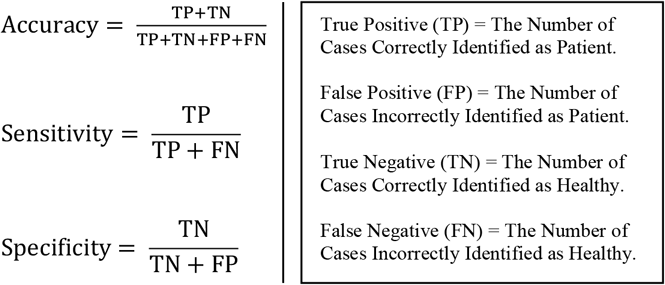

**Graphs:**

## 4. Discussion

### Contributions

Deep convolutional neural networks were used on an optic disc extracted in the first stage to classify images as healthy eyes or glaucomatous eyes. The classification results surpass the previous best methods with a 9.6% relative improvement in terms of area under the curve (AUC) of the receiver operating characteristic (ROC) curve [40 – 44] The approach discussed in this study sets a new state of the art on datasets for localization and also sets the bar for correct localization at IOU > 70. The empirical evidence demonstrates that reporting only AUC for datasets does not portray the true picture of the algorithm’s performance and calls for additional performance metrics to corroborate the results. Thereby, a detailed report of the results, using precision and specificity, was provided in addition to AUC for reliable analysis as well as a thorough future comparison with other methods.

### 4.1 Value Added and Impact

The proposed early diagnostic method for glaucoma using CNNs has displayed efficient results, proving its capability of distinguishing glaucomatous eyes from healthy eyes with accuracy, precision, and specificity. This makes it very helpful in the medical scene as manual observations by ophthalmologists are prone to faults due to clinician bias, fatigue, and human error. This fully automated algorithm is not susceptible to the previously mentioned factors, and offers inexpensive, timely service to those living in remote areas who are unable to access tertiary care setups.

## 5. CONCLUSION

An early diagnostic method using convolutional neural networks is proposed in this study for optic disc localization and classification for glaucoma screening. The optic disc approach was concluded to be contributory to examining the eye for various diseases, namely, ischemic optic neuropathy, compressive optic neuropathy, hereditary optic neuropathy and, most importantly, glaucoma. The disc localization method was employed using convolutional neural networks. This completely eliminates the need for the development of dataset-specific localization methods by providing reliable, robust, and accurate localization across two datasets. The performance of this automated algorithm sets new state-of-the-art results in the two publicly available datasets with Intersection Over Union greater than 70%. The classification of images into diseased and healthy using CNNs has also displayed 98.9% accuracy, 98.8% sensitivity, and 99.1% specificity. Therefore, it is concluded that the methods used in this work are capable of reliably identifying glaucomatous images.

With respect to applications, a glaucoma diagnosis webpage has been designed using the algorithm that was a result of various segmentation and classification training sets. This webpage is accessible to patients wanting to check for glaucoma. The patient can test for glaucoma, and the algorithm either detects the condition and informs the patient ‘to visit the nearest hospital for treatment as soon as possible’ or signals to the patient that ‘the optic nerve is perfectly healthy; no glaucoma detected’. The algorithm is also able to identify if the patient suffers from an optic condition that is widely misdiagnosed as glaucoma.

Additionally, the webpage offers various services, such as providing scientific background about the optic condition and, most importantly, eye treatment training in the form of an interactive video game that patients play while simultaneously improving their focus and strengthening their vision. This webpage approach provides a quick, accessible solution that will allow people to check more frequently for glaucoma, decreasing the chances of it going unnoticed for a long period of time, leading to partial or full blindness. As a result of this approach, glaucoma may eventually stop being the leading cause of blindness worldwide. The glaucoma diagnostic webpage can be found at: [Glaucoma Detector](http://glaucomadiagnosis.com/) (glaucomadiagnosis.com)

## Data Availability

Data was openly available before the manuscript was written.
Sources of data can be found at:
https://www.researchgate.net/publication/345850772_RIM-ONE_DL_A_Unified_Retinal_Image_Database_for_Assessing_Glaucoma_Using_Deep_Learning
https://www.kaggle.com/datasets/lokeshsaipureddi/drishtigs-retina-dataset-for-onh-segmentation

https://www.researchgate.net/publication/345850772_RIM-ONE_DL_A_Unified_Retinal_Image_Database_for_Assessing_Glaucoma_Using_Deep_Learning

https://www.kaggle.com/datasets/lokeshsaipureddi/drishtigs-retina-dataset-for-onh-segmentation

## Notes

### Competing Interest Statement

The authors have declared no competing interest.

### Funding Statement

This study was funded by King Abdullah Medical City

### Author Declarations

DRISHTI-GS Dataset: https://www.kaggle.com/datasets/lokeshsaipureddi/drishtigs-retina-dataset-for-onh-segmentation RIM-One Dataset: https://www.researchgate.net/publication/345850772_RIM-ONE_DL_A_Unified_Retinal_Image_Database_for_Assessing_Glaucoma_Using_Deep_Learning

## References

[1] Liu, Peng, and Ruogu Fang. “Regression and learning with pixel-wise attention for retinal fundus glaucoma segmentation and detection.” arXiv preprint 2001.01815 (2020).

[2] Sivaswamy, Jayanthi, et al. “Drishti-gs: Retinal image dataset for optic nerve head (onh) segmentation.” 2014 IEEE 11th international symposium on biomedical imaging (ISBI). IEEE, 2014.

[3] Fumero, Francisco, et al. “RIM-ONE: An open retinal image database for optic nerve evaluation.” 2011 24th international symposium on computer-based medical systems (CBMS). IEEE, 2011.

[4] McMonnies, Charles W. “Glaucoma history and risk factors.” Journal of optometry 10.2 (2017): 71–78.

[5] Quigley, Harry A. “Schematic Representation of the Anatomy of the Eye.” The New England Journal of Medicine, Massachusetts Medical Society, 15 Apr. 2023, https://www.nejm.org/doi/full/10.1056/nejm199304153281507. Accessed 23 Jan. 2023.

[6] Quigley, Harry A. “Normal and Glaucomatous Optic Disks.” The New England Journal of Medicine, Massachusetts Medical Society, 15 Apr. 2023, https://www.nejm.org/doi/full/10.1056/nejm199304153281507. Accessed 23 Jan. 2023.

[7] Veena, H. N., A. Muruganandham, and T. Senthil Kumaran. “A novel optic disc and optic cup segmentation technique to diagnose glaucoma using deep learning convolutional neural network over retinal fundus images.” Journal of King Saud University-Computer and Information Sciences 34.8 (2022): 6187–6198.

[8] Bussel, Igor I., Gadi Wollstein, and Joel S. Schuman. “OCT for glaucoma diagnosis, screening, and detection of glaucoma progression.” British Journal of Ophthalmology 98. Suppl 2 (2014): ii15–ii19.

[9] Chou, Roger, et al. “Screening for glaucoma in adults: updated evidence report and systematic review for the US Preventive Services Task Force.” JAMA 327.20 (2022): 1998–2012.

[10] Bajwa, Muhammad Naseer, et al. “Two-stage framework for optic disc localization and glaucoma classification in retinal fundus images using deep learning.” BMC medical informatics and decision making 19.1 (2019): 1–16.

[11] Phasuk, Siriporn, et al. “Automated glaucoma screening from retinal fundus image using deep learning.” 2019 41st annual international conference of the IEEE engineering in medicine and biology society (EMBC). IEEE, 2019.

[12] Son, Jaemin, et al. “Development and validation of deep learning models for screening multiple abnormal findings in retinal fundus images.” Ophthalmology 127.1 (2020): 85–94.

[13] Shabbir, Amsa, et al. “Detection of glaucoma using retinal fundus images: A comprehensive review.” Mathematical Biosciences and Engineering 18.3 (2021): 2033–2076.

[14] Elmoufidi, Abdelali, et al. “CNN with Multiple Inputs for Automatic Glaucoma Assessment Using Fundus Images.” International Journal of Image and Graphics (2022): 2350012.

[15] Almazroa, Ahmed, et al. “Retinal fundus images for glaucoma analysis: the RIGA dataset.” Medical Imaging 2018: Imaging Informatics for Healthcare, Research, and Applications. Vol. 10579. SPIE, 2018.

[16] Saravanan, Vijayalakshmi, et al. “Deep learning assisted convolutional auto-encoder framework for glaucoma detection and anterior visual pathway recognition from retinal fundus images.” Journal of Ambient Intelligence and Humanized Computing (2022): 1–11.

[17] Sonti, Kamesh, and Ravindra Dhuli. “Shape and texture-based identification of glaucoma from retinal fundus images.” Biomedical Signal Processing and Control 73 (2022): 103473.

[18] Stein, Joshua D., Anthony P. Khawaja, and Jennifer S. Weizer. “Glaucoma in adults—Screening, diagnosis, and management: A review.” Jama 325.2 (2021): 164–174.

[19] Allison, Karen, Deepkumar Patel, and Omobolanle Alabi. “Epidemiology of glaucoma: the past, present, and predictions for the future.” Cureus 12.11 (2020).

[20] Shoukat, Ayesha, and Shahzad Akbar. “Artificial intelligence techniques for glaucoma detection through retinal images: State of the art.” Artificial Intelligence and Internet of Things (2021): 209–240.

[21] Abdel-Hamid, Lamiaa. “Glaucoma detection from retinal images using statistical and textural wavelet features.” Journal of digital imaging 33.1 (2020): 151–158.

[22] Sarhan, Abdullah, Jon Rokne, and Reda Alhajj. “Glaucoma detection using image processing techniques: A literature review.” Computerized Medical Imaging and Graphics 78 (2019): 101657.

[23] Conlon, Ronan, Hady Saheb, and Iqbal Ike K. Ahmed. “Glaucoma treatment trends: a review.” Canadian Journal of Ophthalmology 52.1 (2017): 114–124.

[24] Schuster, Alexander K., et al. “The diagnosis and treatment of glaucoma.” Deutsches Ärzteblatt International 117.13 (2020): 225.

[25] Bekkers, Amerens, et al. “Microvascular damage assessed by optical coherence tomography angiography for glaucoma diagnosis: a systematic review of the most discriminative regions.” Acta ophthalmologica 98.6 (2020): 537–558.

[26] Liao, WangMin, et al. “Clinical interpretable deep learning model for glaucoma diagnosis.” IEEE journal of biomedical and health informatics 24.5 (2019): 1405–1412.

[27] Öhnell, HannaMaria, Boel Bengtsson, and Anders Heijl. “Making a correct diagnosis of glaucoma: data from the EMGT.” Journal of glaucoma 28.10 (2019): 859.

[28] Li, Fei, et al. “Automatic differentiation of Glaucoma visual field from non-glaucoma visual field using deep convolutional neural network.” BMC medical imaging 18. 1 (2018): 1–7.

[29] Phan, Sang, et al. “Evaluation of deep convolutional neural networks for glaucoma detection.” Japanese journal of ophthalmology

[30] Batista, Francisco José Fumero et al. “Rim-one dl: A unified retinal image database for assessing glaucoma using deep learning.” Image Analysis & Stereology 39.3 (2020): 161–167.

[31] Badawi, Abdulrahman H., et al. “Primary congenital glaucoma: An updated review.” Saudi Journal of Ophthalmology 33.4 (2019): 382–388.

[32] Tran, Jessica H., and Louis R. Pasquale. “A Comparison of Genomic Advances in Exfoliation Syndrome and Primary Open-Angle Glaucoma.” Current Ophthalmology Reports 9.3 (2021): 96–106.

[33] Bui, Trung Thanh, and Jullia A. Rosdahl. “Systematic Review of MIGS and Non-Penetrating Glaucoma Procedures for Uveitic Glaucoma.” Seminars in Ophthalmology. Vol. 37. No. 7-8. Taylor & Francis, 2022.

[34] Flores-Sánchez, Blanca C., and Andrew J. Tatham. “Acute angle closure glaucoma.” British Journal of Hospital Medicine 80.12 (2019): C174–C179.

[35] Scuderi, Gianluca, et al. “Pigment dispersion syndrome and pigmentary glaucoma: a review and update.” International Ophthalmology 39.7 (2019): 1651–1662.

[36] Razeghinejad, Reza, et al. “Pathophysiology and management of glaucoma and ocular hypertension related to trauma.” Survey of Ophthalmology 65.5 (2020): 530–547.

[37] Senthil, Sirisha, et al. “Neovascular glaucoma-A review.” Indian Journal of Ophthalmology 69.3 (2021): 525.

[38] Gillmann, Kevin, et al. “Surgical management of pseudoexfoliative glaucoma: a review of current clinical considerations and surgical outcomes.” Journal of Glaucoma 30.3 (2021): e32–e39.

[39] Ray, Vanita Pathak, Divya P. Rao, and Isha Gulati. “Primary implantation of non-valved glaucoma-drainage-device in sulcus in iridocorneal endothelial syndrome.” International Journal of Ophthalmology 12.11 (2019): 1809.

[40] Abbas, Qaisar. “Glaucoma-deep: detection of glaucoma eye disease on retinal fundus images using deep learning.” International Journal of Advanced Computer Science and Applications 8.6 (2017).

[41] Maheshwari, Shishir, et al. “Automated glaucoma diagnosis using bit-plane slicing and local binary pattern techniques.” Computers in biology and medicine 105 (2019): 72–80.

[42] Ge, Tao, et al. “Seri: A dataset for sub-event relation inference from an encyclopedia.” CCF International Conference on Natural Language Processing and Chinese Computing. Springer, Cham, 2018.

[43] Prasad, Krishna, et al. “Multiple eye disease detection using Deep Neural Network.” TENCON 2019-2019 IEEE Region 10 Conference (TENCON). IEEE, 2019.

[44] Jomar, Deema E., et al. “Risk Factors for Glaucoma Drainage Device Exposure in Children: A Case-Control Study.” American Journal of Ophthalmology 245 (2023): 174–183.

